# Cost burden of type 2 diabetes mellitus (DM) in an urban area of Bangladesh: A hospital-based mix method study

**DOI:** 10.1101/2020.08.03.20167478

**Authors:** Monidipa Saha, Shirmin Bintay Kader, Shafquat Haider Chowdhury, Md. Khaledul Hasan, Mir Nabila Ashraf, Md. Marufur Rahman, Kamrun Nahar Koly

## Abstract

**Background:** Longtime treatment of diabetic patients is an added burden for countries that had low GDP and no health insurance facilities like Bangladesh. Urgent cost assessment and proper strategy needs to be developed to overcome the economic burden of a diabetic patient.

**Methodology:** We have conducted a mixed-methods study in a Bangladesh Institute of Health Sciences (BIHS), a sister concern of BIRDEM hospital between August 2019 to January 2020. Quantitative data was collected by a survey questionnaire from the patients and qualitative data was collected by interviewing five key informants of the facility center.

**Results:** 329 known diabetic patients were enrolled, out of the 70.48% were female and the rest of them were male. The mean age of the respondents was 53.73 years. Nearly 35% of them had a positive family history, and more than 30% of them had both hypertension and dyslipidemia along with DM. The total average cost per year was ∼600USD, which includes drug, investigation, consultancy & transportation costs. Among which drug costs were the most (∼350 USD).We had found that drug cost & investigation cost had a significant associated with increased age, level of education, and the duration of DM. Investigation cost was also associated with positive family history.

**Conclusions:** As there is no insurance coverage in Bangladesh and subsidy are not available, many of the patients from the lower socio-economic group might discontinue the treatment. Further prospective research is needed to develop a proper guideline for policymakers.

**Strength & Limitation:**

- It’s the first mixed method study which discuss about the cost burden of type 2 DM
- KII was performed and perspective about the cost burden of type 2 DM from doctors view was reported.
- Described that treatment of DM is often hampered due to lack of family & workplace support.
- Our only methodological limitation is our sample size, which was smaller than other quantitave studies.

## Introduction

Globally, Non-communicable diseases (NCDs) will become the leading cause of mortality by 2030, and the disease burden will cause severe economic crisis (1). In low and middle-income countries, where about 75.4 % of the total population suffers from diabetes mellitus (DM), yet only 19% of global health expenditure of DM was spent on them (2). As it is a chronic disease, the treatment of DM itself considered a burden for life; it can lead to limit productivity, early retirement, abridged life expectancy, trivial performance for having physical and psychological problems, and even death (3, 4). The alarming issue is non-communicable diseases like diabetes can particularly affect the economically deprived population, who are more prone to disease complications and mortality (5).

According to The International Diabetes Federation (IDF), the global epidemic of diabetes will increase by 48% from 425 to 629 million by 2045 (6). A projection estimated that the South-East Asia Region will experience an increase in health expenditure on diabetes in the next decades, reaching USD 10.1 billion in 2030 and USD 12.3 billion in 2045 (2). An economic analysis reported that individuals with diabetes have a 2.3 times higher average of medical expenditures than an individual without the condition, which clearly states the financial burden on an individual. (3). Adults with diabetes in India, Bangladesh, and Sri Lanka make up 98.9% of the total adult population with diabetes in the region. Bangladesh is among the top 10 countries, with 8.4 million people living with diabetics (2). Studies conducted among a small-scale population in Bangladesh showed an increasing trend of diabetes prevalence in rural and urban communities (7). A diabetic patient in Bangladesh pays a considerable amount for the treatment due to lack of access to adequate medical services, health facilities, lack of resources compared to the other developing countries (8). Average annual spending per person was found in USD 64 in Bangladesh (2).

Lack of health awareness and hardship in accessing health care services in Bangladesh are the tip of the iceberg in controlling DM; there are other factors involved in keeping the disease under control. Unfortunately, relevant literature about out-pocket expenditure associated with DM is limited due to the absence of large-scale studies in Bangladesh. Thus, the actual scenario of prevalence and cost burden of DM remains unexplored in Bangladesh. The current study provides a snapshot of the out-pocket expenditure of DM among the patients receiving care from the Keranigonj Healthcare Centre, Bangladesh, which might be able to draw an outline of recommendations for healthcare policymakers. We have also explored the perception of the doctors to understand the out-pocket expenditure of type 2 DM from the health care providers’ perspective.

## Methods and Materials

### Study Design

We have conducted a cross-sectional study with a mix-method approach in a primary non-government healthcare center located in a suburban area to find out the cost burden of type 2 diabetes mellitus (DM) in Bangladesh. We conducted both the quantitative and qualitative surveys to evaluate the direct and indirect costs involved in treating patients with type 2 DM.

### Study Site, size & population

For the cross-sectional survey, the study participants were enrolled from the patients who visited Keranigonj Healthcare Centre (KHC), which has an outpatient unit of the Bangladesh Institute of Health Sciences (BIHS). BIHS is a sister concern of Bangladesh Institute of Research and Rehabilitation in Diabetes, Endocrine, and Metabolic Disorders (BIRDEM). All the diabetes patients who visited the KHC had a health check-up card, which was provided by BIRDEM. We have included 329 patients with Type II diabetes in between August 2019 to January 2020 purposively. We excluded the patients who had a history of organ failure (i.e., Kidney, Heart) and who was critically ill at that moment.

For the qualitative survey, Doctors from the outdoor department of the KHC invited as a key informant to perform KII (Key Informant Interview). Five outdoor senior medical officers were interviewed to conduct the Key Informant Interview.

### Data collection tools

#### Survey questionnaire

After taking consent from the patients, we have conducted a face-to-face interview by using a semi-structured questionnaire. The survey tool included a component to measure the direct and indirect costs subjectively alongside the socio-demographic status of the participants. The direct costs considered the price directly related to treatment (i.e., drug, consultancy fees, and investigation fees), and other expenditures such as transport costs were considered as the indirect cost. We have also collected information about the pre-existing co-morbidity and the current medication histories of the patients. The questionnaire was prepared in English and then was translated into Bangla and has been pre-tested before the survey.

#### Key informant interviews

A total of five KIIs were performed by developing an interview guideline after extensive literature review, (supplementary text) interview was done at the hospital settings. Each session was conducted for 40-60 minutes by the trained researcher, and the entire session was designed to allow spontaneous discussion. The discussions were audio-recorded with proper consent, and the interviewer also kept field notes for future reference in analysis. Once data saturation was obtained, we stopped collecting information for the qualitative part.

### Data processing and analysis

We used Stata 13 (College Station, Texas 77845 USA) to summarize and analyze the descriptive statistics. To find out the associating factors related to cost burden at different age groups, we did one way ANOVA. To find out the relation of different costs with characteristics, we considered Kruskal-Wallis and Wilcoxon Rank Sum test. We have considered total drug cost, investigation cost, and consultancy cost separately. We categorized the data to perform simple and multiple median regressions to determine the variables responsible for out-pocket expenditure and visualized the result by using MedCalc Statistical Software version 19.1.7.

For qualitative analysis, we transcribed the KIIs in Bangla and translated them into English. Once transcription of the KIIs was completed, we read the data thoroughly to get an idea about the data pattern and checking the consistency. Then a codebook was developed for data coding. We coded the data using MS excel according to the codebook. We prepared a matrix table once coding was done, and a summary was developed from the data matrix. We used thematic analysis approach to analyze the qualitative part.

### Ethical issues

The Approval was obtained from The Ethical Review Committee (ERC) of the American International University Bangladesh (AIUB). Written permission was taken from the Keranigonj Healthcare Centre administration as well. Both verbal and informed written consent in Bangla was taken from study subjects.

## Results

### Sample demographic

A total of 329 adults were enrolled in our study, where 70.48% (232) were female, and 29.48% (97) were male. The average monthly income was 22074.47±14958.17. The mean age of the participants was 53.73 years, where the maximum age was 85 years, and the minimum was 21 years. The average duration of having DM was 5.46± 3.90 years.72.04% of them were aged above 45 years. Among the participants, 92.10% were Muslim, 6.08% were Hindu, and 1.82% were Buddhist. 36.48% of the participants have completed secondary education, 24.62% have completed higher secondary education, 27.35% have completed a bachelor’s degree, and only 11.55 % have completed their master’s degrees. 44.07% of the participants were employed. 34.35% of the participants had a positive family history of type-2 DM, and 31.61% had both HTN & dyslipidemia along with DM. Only fewer (17.33%) had none of hypertension and dyslipidemia. (Table: 1)

**Table 1:** Socio-demographic data:

| **Variable:** |  | **Mean/ N (%)** |
| --- | --- | --- |
| Age |  | 53.73±12.61yrs |
|  | Below 45yrs | 92(27.96%) |
|  | Above 45yrs | 237(72.04%) |
| Sex |  |  |
|  | Male | 97 (29.48) |
|  | Female | 232 (70.48) |
| Monthly income |  | 22074.47±14958.17 |
| Religion: |  |  |
|  | Islam | 303 (92.10) |
|  | Hindu | 20 (6.08) |
|  | Buddhist | 6 (1.82) |
| Education |  |  |
|  | Completed Secondary Education | 120 (36.48) |
|  | Completed Higher Secondary | 81 (24.62) |
|  | Completed Bachelor Degree | 90 (27.35) |
|  | Completed Master Degree | 38 (11.55) |
| Occupation |  |  |
|  | Unemployed | 184 (55.93) |
|  | Employed | 145 (44.07) |
| Duration of the disease |  | 5.46± 3.90 yrs |
| Family History of DM |  |  |
|  | Yes | 113 (34.35 %) |
|  | No | 167 (41.64%) |
|  | Unknown | 49 (14.89%) |
| Co-morbid Conditions |  |  |
|  | Hypertension | 117 (34.35 %) |
|  | Dyslipidaemia | 51 (15.50%) |
|  | Hypertension & Dyslipidaemia | 104 (31.61%) |
|  | Absent | 57(17.33%) |

### The cost difference in different age group

The total cost of type-2 DM varied between different age groups. We found that the average monthly total cost per person suffering from type II diabetes was 4083.53 BDT (yearly ∼600USD). Here the total cost includes drug cost, consultancy paid, investigation cost, and transportation cost. The monthly drug cost per person was2518.39 BDT (yearly∼350 USD), consultancy paid per month 254.74 BDT (yearly ∼35 USD), spending behind the investigation was1145.53 BDT (yearly∼ 160 USD) and transportation cost was 164.86 BDT (yearly∼24 USD). Drug cost, investigation cost, and total cost of type II diabetes increased with age and found statistically significant. (p<0.001) (Figure: 1). The highest average yearly total cost (4863.47BDT) of treatment was observed in the 51-60 years group, where the lowest (2241.11BDT) was reported in the younger age group (21-30 years). A similar scenario was found in cost spears for drugs also. Transportation cost and consultancy cost was more or less similar for different age groups and also statistically non-significant. We noticed that the reason behind the cost burden was primarily the price of the drugs, investigation costs, transport, and consultancy costs. (Table: 2)

**Figure 1:**
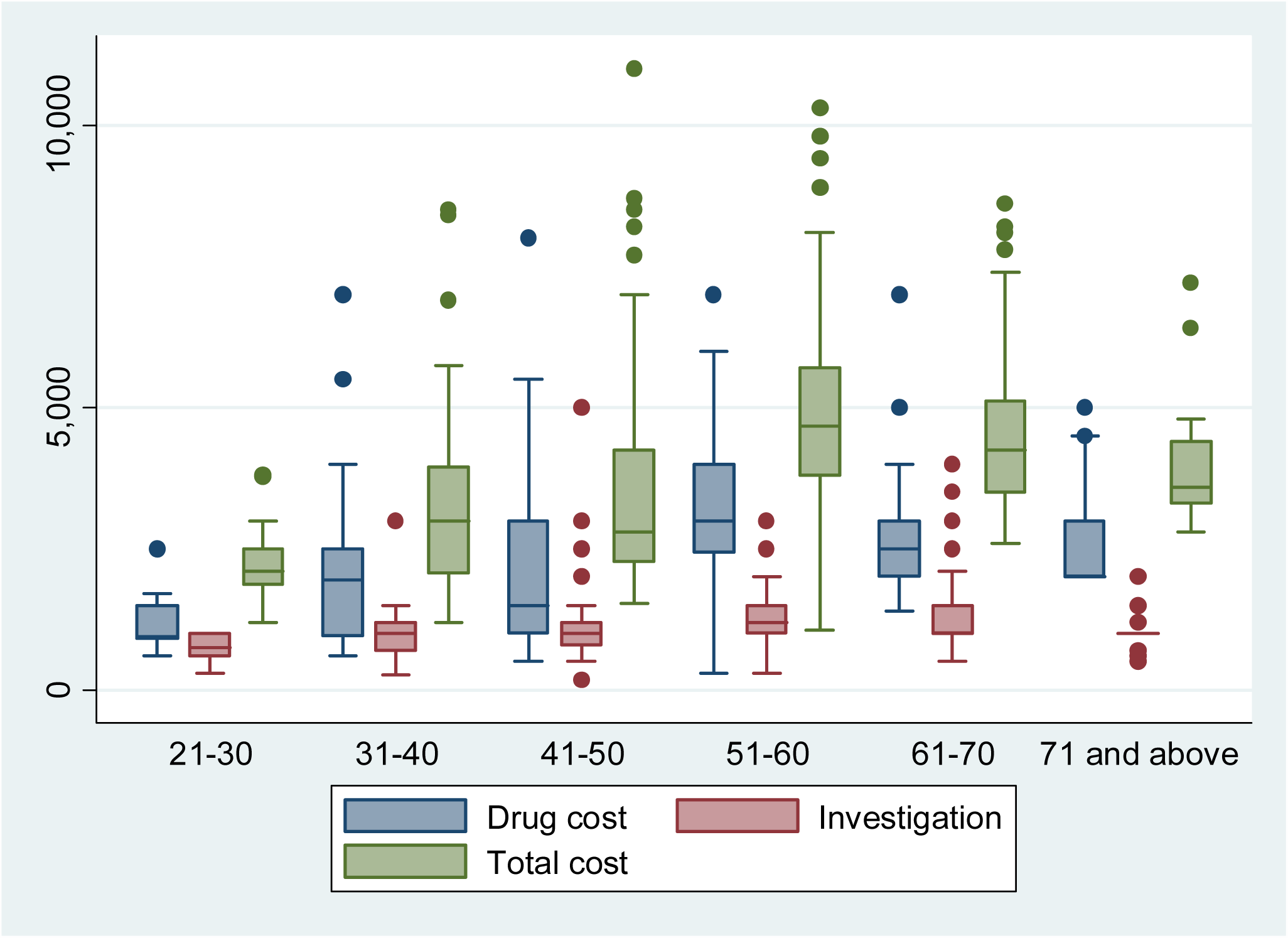
Cost distribution of different age groups.

**Table 2:** Mean Costs in different age group:

| Cost | Mean | 21-30yrs | 31-40 | 41-50 | 51-60 | 61-70 | 71 & above | P-Value |
| --- | --- | --- | --- | --- | --- | --- | --- | --- |
| **Total Cost** | 4083.53 | 2241.11 | 3368.61 | 3555.66 | 4863.47 | 4514.56 | 3936.19 | 0.000 |
| **Drug Cost** | 2518.39 | 1183.33 | 2004.17 | 2038.55 | 3213.04 | 2743.04 | 2552.38 | 0.000 |
| **Consultancy** | 254.74 | 200 | 247.22 | 242.17 | 245.11 | 304.56 | 219.05 | 0.161 |
| **Investigation** | 1145.53 | 744.44 | 975.28 | 1102.05 | 1228.26 | 1301.26 | 1004.76 | 0.0003 |
| **Transportation** | 164.86 | 113.33 | 141.94 | 172.89 | 177.07 | 165.70 | 160 | 0.701 |

### Relationship between mean cost and Patient’s Characteristics

The relationship between different cost components and characteristics of the patients were analyzed by Wilcoxon Rank Sum and Kruskal-Wallis Test. The overall total cost, along with drug & investigation costs, were significantly correlated with age, employment status, presence of co-morbidity & family status. Gender had no significant role in cost difference for investigation, drug, and total costs. The patient who suffered from type 2 DM more than 5 years had significantly higher drug costs compare to those suffered less than 5 years. Both the investigation & drug cost were higher in the patient had no family history of type 2 DM than with a family history. Co-morbidities, especially dyslipidemia, enhanced the treatment cost, where investigation cost was shown less variance (table: 3).

**Table 3:**
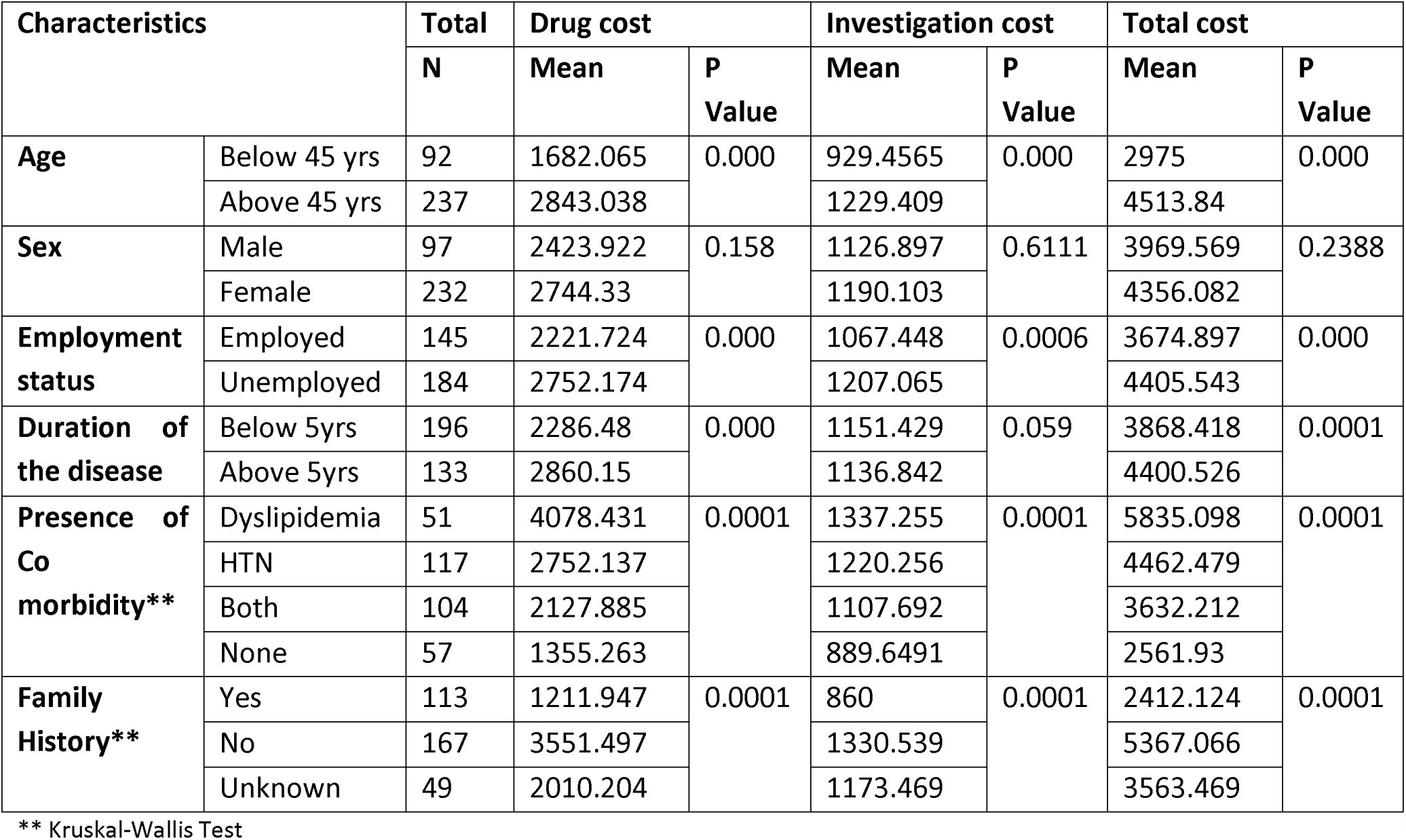
Relationship between cost and patient’s characteristics:

### Determination of out-pocket expenditure considering confounders

We categorized the costs into different scales depends on their scale. Total cost, drug cost, investigation cost, and consultancy cost were categorized into two groups below and above 3500, 2500, 1000, and 200 BDT, respectively. Simple and multiple logistic regression was performed, results were presented by plotting into forest plot. (Detail results were not provided here). From the analysis, we found that Age ranges, sex, education, occupation, duration of type 2 DM, family history with DM, and use of OAD or Insulin drugs were not associated with the total costs. When we consider separate costs, we have seen that who had educational qualification above HSC they spend around 4 times and 6 times more than who had up to HSC education in drug and consultancy cost respectively.(p<0.0001). Respondents of more than 45 years old and had a family history of type 2 DM were more likely to spend double for diagnosis and disease investigation (AOR 2.1529, p=0.0355 & AOR 1.8474, p=0.01 respectively). (Figure 2)

**Figure 2:**
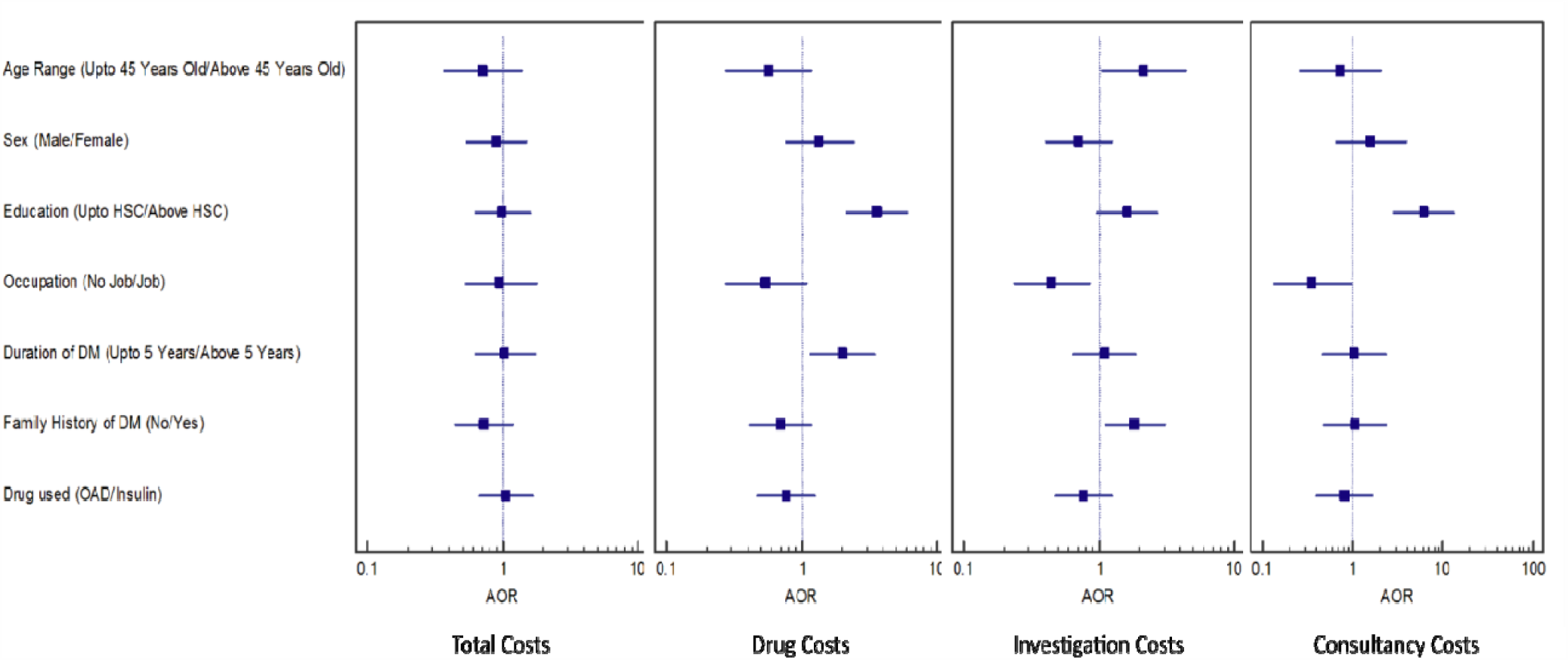
Forest plot describing the factors responsible for economic burden.

## Doctors’ perspective regarding the cost burden of DM (findings from KIIs)

### Socio-economic characteristics of the doctors

All the five doctors, interviewed as a key informant, were senior medical officers by designation. 3 of them were female, and the rest of the doctors were male. 2 of 5 doctors were from the age range of 35-40 years, one of them was from 41-45 years, and the rest of the 2 doctors were from the age range of 46-50 years. The doctors were well experienced as most of the doctors (3 of 5) had more than 15 years of work experience (2 female, 1 male). The others had a work experience of 8 years (1 male, 1 female).

### Socio-economic characteristics of the patients

The majority of the doctors (3 of 5) mentioned that the patients were mostly from the low socio-economic group, whereas the rest of the doctors (2 of 5) reported that lower to a higher middle-income group of patients visited them for diabetes treatment. One of the doctors thinks that diabetes is now a common disease for all the socio-economic groups. He added:

*“Unfortunately, it seems diabetes is no more a disease of the high socio-income group as it was perceived before. Most of the newly diagnosed diabetic cases are from poor economic backgrounds”.*

### Lack of family support

Female patients miss their follow up to more than male patients. According to the view of the doctors, more than 80% of the patients, aged 50 years or above miss their regular follow up, and most of them are females who are from lower socio income groups. Female patients are often neglected by their husbands or family members and treated as a burden to the family. A high percentage of female garment workers also miss their follow up as they do not want to lose their working hours. As one of the doctors reported:

*“A good number of patients complain that they are not being able to make their family happy due to this disease. The female patients often complain that their husbands are always shouting at them as they cannot work properly due to diabetes”.*

### Financial Barrier

Financial constrain was the main barrier for diabetic treatment as reported by all doctors. The high price of diabetic-related drugs was a burden for the lower-income groups. Therefore, medication adherence was low in this group, as reported by the doctors. All the doctors mentioned that about 85% of patients fail to buy medicine due to the high cost, which is beyond their affordability. The patients often complain about the high price of the drugs. The doctors also added that, in terms of diabetic check-up and investigation, 80% of patients fail to do their regular check-up due to the high price of tests. Majority of the doctors (4 of 5) mentioned that the elderly age group of patients mainly complains about the cost of treatment whereas one of the doctors reported that both mid and elderly age group of patients complains about the high cost of diabetes treatment. As one of the doctors added:

*“Patients complain mostly about the high price of medicine and investigation. They say doctors should prescribe low-cost medicine”.*

### Cost of the drugs

All of the doctors mentioned that most of the patients prefer oral drugs over insulin, as the patients think that insulin is more costly than oral medication. The doctors also reported that patients think that they do not have enough facilities to preserve insulin. Also, most of the patients are afraid of the injection. So they prefer oral drugs in most cases. As one of the doctors added:

*“All of the patients want an oral drug. Most of them are afraid of taking an injection, and they also think insulin is costly”.*

Among the lower-income groups, there was a tendency of not following the diabetic food chart as reported by the doctors. The patients are also not aware of maintaining the food chart as they cannot afford to buy foods only for diabetes management. Most of the doctors (3 of 5) mentioned that, as the poor patients do not follow the diabetic food chart, they do not think that their food cost has increased due to diabetes. One of the doctors said:

*“No, they don’t think that food cost has increased. Only a few from good educational background think they need to spend on food”.*

## Discussions

This cross-sectional study was conducted among 329 patients with type 2 DM who had visited Keranigonj Healthcare Centre (KHC), which has an outpatient unit of Bangladesh Institute of Health Sciences (BIHS). The aim of the study was to find out the out-pocket expenditure of DM among the patients receiving care from the Keranigonj Healthcare Centre, Bangladesh, and also to explore the perception of the doctors regarding the cost burden of DM from health care provider’s perspective.

Among the 329 patients, around 70.48% (232) were female, and 29.48% (97) were male. Though type 2 DM is more common in males throughout the world, however, in our study, we found the prevalence was higher among females compared to males. The result is similar to other studies conducted in Bangladesh, which had also reported that diabetes was more prevalent among women than men (12, 13). The mean age of our study participants was 53.73±12.61 that might be due to increasing life expectancy in Bangladesh, which is similar to the previous study conducted in Bangladesh, India, and Ghana (5, 14, 15). Often we have seen that the treatment cost of Diabetes with other co-morbidities such as hypertension or the presence of dyslipidemia are more expensive than Diabetes without any co-morbidities (5). In our study, we found that 31.61% of the study participants had both HTN & dyslipidemia, which is similar to the findings from Vietnam (15).

The economic aspect and the cost burden of type 2 DM are a snapshot of the country’s scenario. So, to maintain sustainable development, it’s essential to understand both beneficiary and influencers’ factors that are responsible for the cost burden of the disease. Type 2 DM is a lifelong disease that has an essential role in the global cost burden. (9). The disease is considered to affect the economic weight for both individuals and, at the same time, collectively for the as a nation as well.

We have considered drug cost, Consultancy fee, and Investigation cost, transport cost, and accumulated all of those to find the total cost burden of Type 2 Diabetes. To avoid biases, we have collected monthly expenditure from respondents. For comparing our study result with other studies, we have converted the yearly average cost from BDT to USD. The total annual cost for the treatment of type two diabetic patients was ∼600USD, which is in between two findings that were conducted in Bangladesh previously (14, 16). In a developing country like Bangladesh, where the GDP is 2,173 U.S. dollars per person in 2020 (2), the average annual cost for each person with type two DM clearly a burden to them. It indicates that people suffering from type 2 DM spend nearly 28% of their per capita income.

This is lower than previous findings that are notified by Afroz et al. (16). When we considered age distribution, we saw that 51-60 years of age group patients expensed more than any other group in every section (i.e., total, direct, and indirect costs), which is similar to a previous study conducted in Bangladesh (17). In our study, we have seen that patients expended most for drugs and investigations; we tried to correlate the responsible factors for total treatment, drug, and investigation costs similar to Samuel et al. study (5). We found that gender is not associated with the total cost and direct costs related to treatment. However, from KII, we came to know a different aspect regarding this. One of the key informants mentioned that though the cost burden is non-significant to sex, in the context of Bangladesh, where most of the women are financially dependent on their husband had to struggle twice. Where the age ranges, duration of diabetes, family history of diabetes, and presence of co-morbidities were correlated with the total & direct cost of the disease, similar to the previous study findings (5, 14, 16). In logistic regression, we have seen that patients who went for graduation were positively correlated with increased drug and consultancy costs. This is similar to the findings of a previous study held in Bangladesh (14).

The limitation of our study was we have interviewed only 329 adults, where most of the studies were conducted on large sample size (18). However, our results are similar to the other studies conducted previously in Bangladesh. Another limitation of our study is, it fails to conduct sensitivity analysis between income and expenditure of the participants; we have compared the out-pocket expenditure with per capita GDP instead. The strength of our study is we have conducted five KIIs, which gives us a clear view of the cost burden scenario faced by diabetic patients.

## Conclusions

The rise in the prevalence rate of type 2 diabetes in Bangladesh where out of pocket health expenditure is more and health insurance coverage doesn’t reach the common people made us interested to know how much the cost of diabetes treatment is making people suffer. For this study, a good number of independent variables are considered, keeping in mind the studies regarding this done in other parts of the world. Though this study doesn’t include the indirect cost, especially cost due to productivity loss. Still, it gives a clear idea about the components of the direct costs. The cost calculated being done as per person per year gives an exact idea about the out-pocket expenditure faced by each suffering from diabetes. It showed how tough it is for a low and mid-income group of people to bear the expenses for diabetes patients. Though the cost calculated is much lesser than the cost found in other countries, especially the Western countries of the world, we need to keep in our inflation rate, GDP, and expenditure in the health sector by the government is much lesser than the western countries. At the same time, as diabetes is no longer a disease for the rich and to add on it, we don’t have a formulated health insurance system in the country makes it almost impossible for the people of the low and mid-income groups to continue with the treatment. This is reflected in the interviews given by the doctors. And though we talk about women empowerment, still women are the most sufferers as their husbands don’t want to pay the extra cost of treatment. This was also stated by the doctors who were interviewed. One more thing needed to be addressed that most of our doctors don’t follow the national guideline for treating a diabetes patient for these chances of polypharmacy increases, which ultimately increased the drug cost. It’s important to know that though none of the variables have any significance on the total cost, individual components of the total cost vary a lot due to the independent variables. It’s also crucial to carry out a nationwide Cost of Illness (COI) study for diabetic patients to understand whether treatment cost varies as per geographical location, which has been reported in different Indian studies. The inflation rate also plays an important role in increasing the COI year by year. So keeping that in mind, a formula needed to establish to understand what will be the COI of 5 or 10 years down the line. The cost of illness and expenditure due to illness are two different things. The cost of illness and expenditure behind illness may not be the same in countries like Bangladesh. Here expenditure like your buying capacity, whereas COI is the amount required if treatment of disease needed to be done as per guideline. The study of the cost of illness gives you an idea to formulate a proper policy for the budget allotment in the health sectors. Diabetes, with its high mortality and high cost of treatment, will always remain a burden unless prevention and standardized health insurance policy are not adopted on an urgent basis.

## Recommendations

i. Cost-effective strategies should be planned to manage and prevent diabetes
ii. A proper insurance policy should be developed, which will include special facilities for diabetes patients.
iii. The government should provide more subsidies on overall drugs used for the treatment of Diabetes.
iv. Nationwide diabetic registration program should be ensured to make sure the target population receives both medical and financial help on time.
v. Special monitoring cell needed to be set up to control the treatment price of diabetes patients in the private sector.
vi. Especially more attention and subsidy should be given to those pharmaceuticals who are producing insulin.
vii. Early diagnosis of diabetes is crucial to minimize the adverse disease outcome, which can be done raising awareness and breaking the social stigma, especially for women in Bangladesh.

## Data Availability

Data is available on reasonable request

## Acknowledgment

We would like to thank AIUB for giving the ethical clearance and Bangladesh Institute of Health Sciences (BIHS) for allowing us to collect data. Three of the authors (SBK, MKH and KNK) are currently employees of icddr,b. icddr,b is grateful to the Governments of Bangladesh, Canada, Sweden, and the UK for providing core/unrestricted support.

## Conflict of Interest & funding

None

## Notes

### Competing Interest Statement

The authors have declared no competing interest.

### Funding Statement

It was an unfunded research work

### Author Declarations

Ethical clearance was taken from ethical review committee of American Internation University-Bangladesh

